# Networked SIRS model with Kalman filter state estimation for epidemic monitoring in Europe

**DOI:** 10.1101/2025.02.06.25321780

**Authors:** Atte Aalto, Daniele Proverbio, Giulia Giordano, Alexander Skupin, Jorge Gonçalves

## Abstract

**Background:** Metapopulation models, which consider epidemic spread across interconnected regions, can provide more accurate epidemic predictions with respect to country-specific models. Still, their added complexity and data requirements raise questions about their tangible benefits over simpler, localized models.

**Aim:** Our goal is to develop and validate networked metapopulation SIRS models, integrated with an Extended Kalman Filter (EKF), for predicting influenza-like illness (ILI) across Europe, which enables accurate forecasts, missing data imputation, and actionable insights.

**Methods:** We constructed two different metapopulation SIRS models: a detailed network-based model, including inter-country travel dynamics, and a simpler mean-field model, aggregating average regional data. Both were calibrated based on decade-long data of European mobility and ILI incidence, using EKF to estimate disease dynamics and forecast epidemic progression. The forecasting performance was benchmarked against isolated country-specific models.

**Results:** Network models outperformed isolated models in forecasting ILI progression, particularly during critical periods such as wave onsets and peaks, and maintained reliability during COVID-19-affected seasons. The full network model provided up to 25% improvement in peak predictions and demonstrated robustness in imputing missing data, even when up to 40% of the input data was unavailable. The models are fully interpretable and align with epidemiological dynamics across borders.

**Conclusion:** Our findings unveil the advantages of metapopulation models for epidemic forecasting in interconnected regions. Our framework, combining network models with EKF, offers improved accuracy, resilience to missing data, and enhanced interpretability. Our methodology provides a versatile tool for global public health applications, adaptable to other diseases and geographic scales.

## 1 Introduction

Modern epidemiology increasingly relies on mathematical models to integrate empirical data, perform advanced analytics, and provide quantitative predictions that help forecast and control the spread of infectious diseases [1]. These models not only predict epidemic progression and future scenarios, but also support the development of early warning systems [2], the assessment of the impact and effectiveness of restrictions [3], and the estimation of unreported cases [4] or cryptic transmission [5]. Hence, advancing forecasting methodologies is crucial to enable timely and effective public health responses, as testified by initiatives like Influcast [6], aimed at developing and testing cutting-edge approaches for influenza-like illnesses (ILI).

Traditional epidemiological models often focus on individual countries, without capturing the broader dynamics of interconnected regions. In contrast, metapopulation models [7, 8], based on the theory of epidemic spreading on complex networks [9, 10, 11] and multi-patch mathematical models [12, 13], can predict the spatiotemporal progression of infectious diseases across large interconnected areas by incorporating mobility networks [14, 15].

Recent studies have successfully employed network-based metapopulation models to forecast real-world epidemic dynamics, including COVID-19 across Italian regions [16, 17], influenza across U.S. states [18, 19], and dengue in China’s Guangdong province [20]. Despite the significant potential of metapopulation models, a critical question persists: do they offer substantial improvement in forecasting accuracy compared to isolated models [21]?

Here, we investigate whether a network-based metapopulation model improves epidemic forecasts for ILI over multiple seasons in Europe. Forecasting ILI is essential for preparedness of healthcare systems and resource allocation. The European Centre for Disease Prevention and Control (ECDC) has a long tradition of data sharing and model evaluation [22]: initiatives such as Respicast [23] provide a platform for integrating and benchmarking diverse forecasting models. We develop novel metapopulation models and evaluate their performance against corresponding country-specific models. These country-specific models were among the top-performing methods in the Respicast dashboard during the season 2023–2024.

Our approach uses ten years of weekly ILI incidence and empirical data on European mobility to construct two models of increasing complexity: a mean-field model focuses on the epidemic evolution within a country and the average incidence in Europe, requiring relatively few data; a detailed network-based model accounts for inter-country travel dynamics and requires additional data on the network structure of mobility between countries. To estimate and predict disease dynamics, both models use an Extended Kalman Filter (EKF), an approach already used in epidemiological studies, such as [4, 18, 19, 20, 21, 24].

Our results reveal the benefits of metapopulation models for epidemic forecasting in interconnected regions, offering probabilistic forecasts and scenario modelling at a continental scale. Our findings demonstrate that augmenting localized information with network-based data improves prediction accuracy and resilience to missing data. Moreover, our framework can be adapted to other diseases and geographic scales, and thus offers a versatile tool for global public health applications.

## 2 Materials and methods

### 2.1 Network generation using mobility data

To construct the metapopulation model, we first developed a mobility network representing population flows between European countries; see Figure 1a. The network was built using publicly accessible data sources, ensuring consistency and comprehensive coverage. Alternatively, the network could be inferred from epidemic data, but this has been shown to be a severely ill-posed problem [25]. Each country was modeled as a node, with links representing three key types of travel flows: air and ferry travel, cross-border commuting, and other forms of land-based movement between neighbouring countries. These components account for diverse modes of mobility, providing a realistic depiction of cross-border interactions. Here, we provide an overview of the network structure; further details are in Supplement S1.

**Figure 1:**
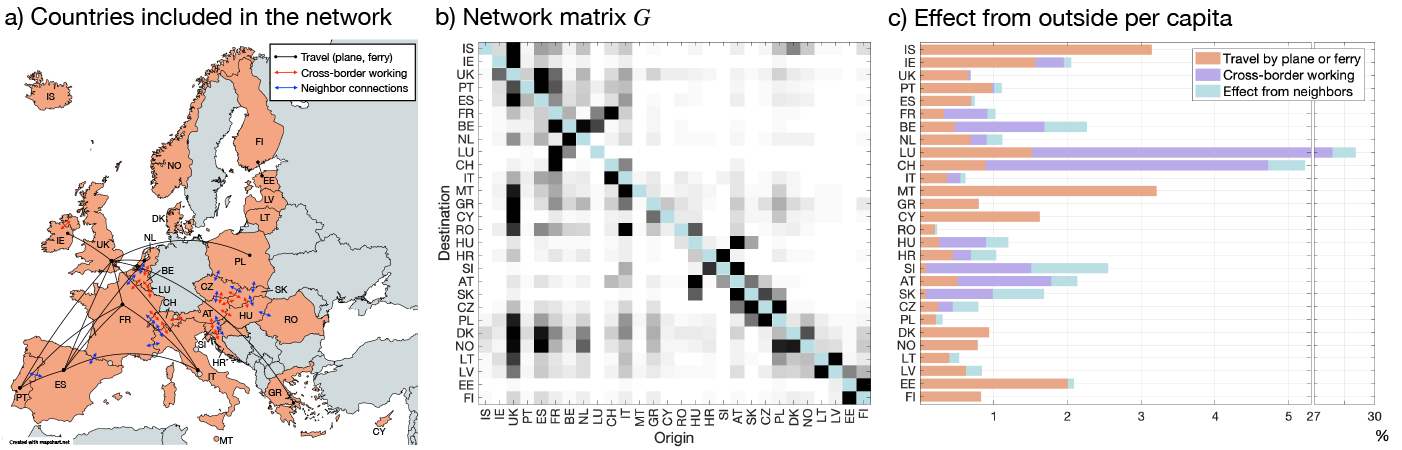
**a:** The countries included in the network are highlighted in light red. The 15 busiest travel connections (by plane or ferry on any route between the indicated countries) are indicated with black lines (*>*10,000 passengers/day/direction). Red arrows show the 15 busiest cross-border worker flows (*>*3,000 workers), and blue arrows show the 15 strongest neighbour connections of other types. **b:** Visualization of the network matrix *G*. Each row has been normalized to have maximum value one. Darker color indicates stronger effect. **c:** The effect from the outside for each country, calculated as the row sums of the network matrices *1*_1_*G*^(1)^, *1*_1_*G*^(2)^, and *1*_2_*G*^(3)^ shown as percentages of the respective countries’ populations. Only effects from other countries included in the network are considered.

#### Air and ferry travel

Data on air travel were sourced from the Eurostat Data Browser (see the “Data sources” section below). Ferry traffic statistics were obtained from the Finnish Port Association and the UK Department for Transport. The air travel data, available quarterly, were averaged to estimate annual levels. While COVID-19 disrupted air and ferry travel patterns, we used a fixed network for the entire period based on either pre- or post-pandemic data.

#### Cross-border commuting

Cross-border worker flows were extracted from reports by the European Commission’s Directorate-General for Employment, Social Affairs and Inclusion [26] and the Economic and Social Research Institute [27]. These flows were calculated based on daily commuting patterns via road and trains, adjusted to reflect weekends, public holidays, leave periods, and travel in both directions. Commuter flows between non-neighbouring regions were excluded, as they are typically captured within the air and ferry travel data.

#### Land-based movement

For neighbouring countries, land-based movement was estimated using border region population densities and border lengths, using the NUTS 3 regional classification (https://doi.org/10.2785/714519). This approach accounts for informal and unregistered travel, which is not available in public databases. While this method provides reasonable approximations, future improvements could incorporate more granular datasets to refine these estimates.

#### Network structure and parameters

Our network matrix integrates the three mobility components, weighted by tuning parameters γ_1_ and γ_2_, to account for the relative contributions of different forms of travel. The population fluxes corresponding to air-and-ferry and commuting flows, denoted by matrices *G*^(1)^ and *G*^(2)^, were combined and scaled by γ_1_, while the land-based movement component, denoted by *G*^(3)^, was scaled by γ_2_. The resulting network matrix

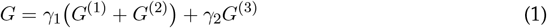

captures the influence of mobility on epidemic dynamics, enabling detailed modelling of interconnected populations. The complete network is visualized in Figure 1b as the heatmap of the row-wise normalized adjacency matrix *G*_*i,j*_*/* max(*G*_*i,·*_) between countries of origin and destination. The network captures detailed inter-country interactions. For example, the strong connectivity between Austria and Slovakia reflects their shared borders and economic ties, which influence mutual epidemic progression.

#### Regional impacts of the network

In almost all countries, the contribution of traveling people to the spread of ILI diseases is estimated to be less than 5% (Figure 1c). An exception is Luxembourg, where cross-border workers represent an important fraction of the effective population. For island countries, or those where the major urban areas are close to shores (*e.g*., Iceland, UK, or Finland), the largest effect is due to air and ferry travel.

By combining diverse mobility data sources into a unified framework, our network generation approach provides a foundational framework to simulate disease spread across Europe. The methodology is flexible and can be extended to other countries or regions as new data become available.

### 2.2 Incidence data

#### Data sources and preprocessing

Influenza-like illness (ILI) incidence data were obtained from the “Respicast – European respiratory diseases forecasting hub” project [23], which consolidates data from the ECDC’s European Respiratory Virus Surveillance Summary (ERVISS) and the World Health Organization’s (WHO) FluID global influenza program. The dataset spans ten epidemic seasons, from week 40 of 2014 to week 16 of 2024, providing a broad temporal and spatial overview of ILI progression across Europe. Reported incidences are provided as weekly cases per 100,000 inhabitants. To enable integration into the metapopulation model, these incidences were scaled to estimate the actual number of detected infections for each country. For the United Kingdom, data were aggregated from England, Wales, Scotland, and Northern Ireland to align with available mobility data, which were reported for the whole UK.

#### Data characteristics

Figure 2 illustrates the population-weighted average ILI incidence across Europe over the study period, highlighting wave onsets and peaks. These critical periods were used to evaluate forecasting performance in our analysis. The dataset provides a comprehensive basis for model calibration and evaluation, offering insight into the seasonal dynamics of ILI at a continental scale.

**Figure 2:**
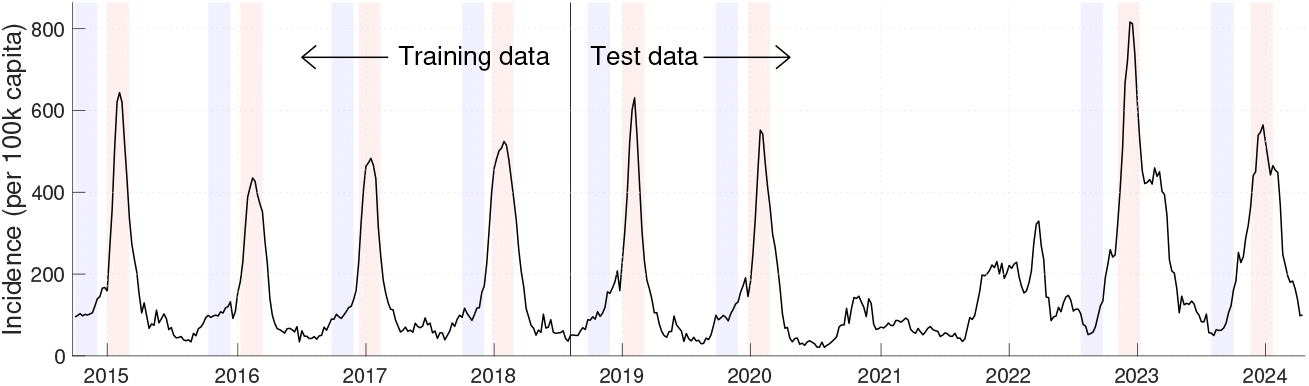
Population-weighted average incidence of influenza-like illness (ILI) across Europe over ten epidemic seasons (2014–2024). Blue-shaded areas indicate wave onset periods, while red-shaded areas mark epidemic peaks. These critical periods were used for evaluating fore-casting performance in subsequent analyses. The data were aggregated from weekly reports and adjusted for population differences across reporting countries. The figure highlights the seasonal dynamics of ILI and the variation in timing and intensity of waves over the study period.

**Figure 3:**
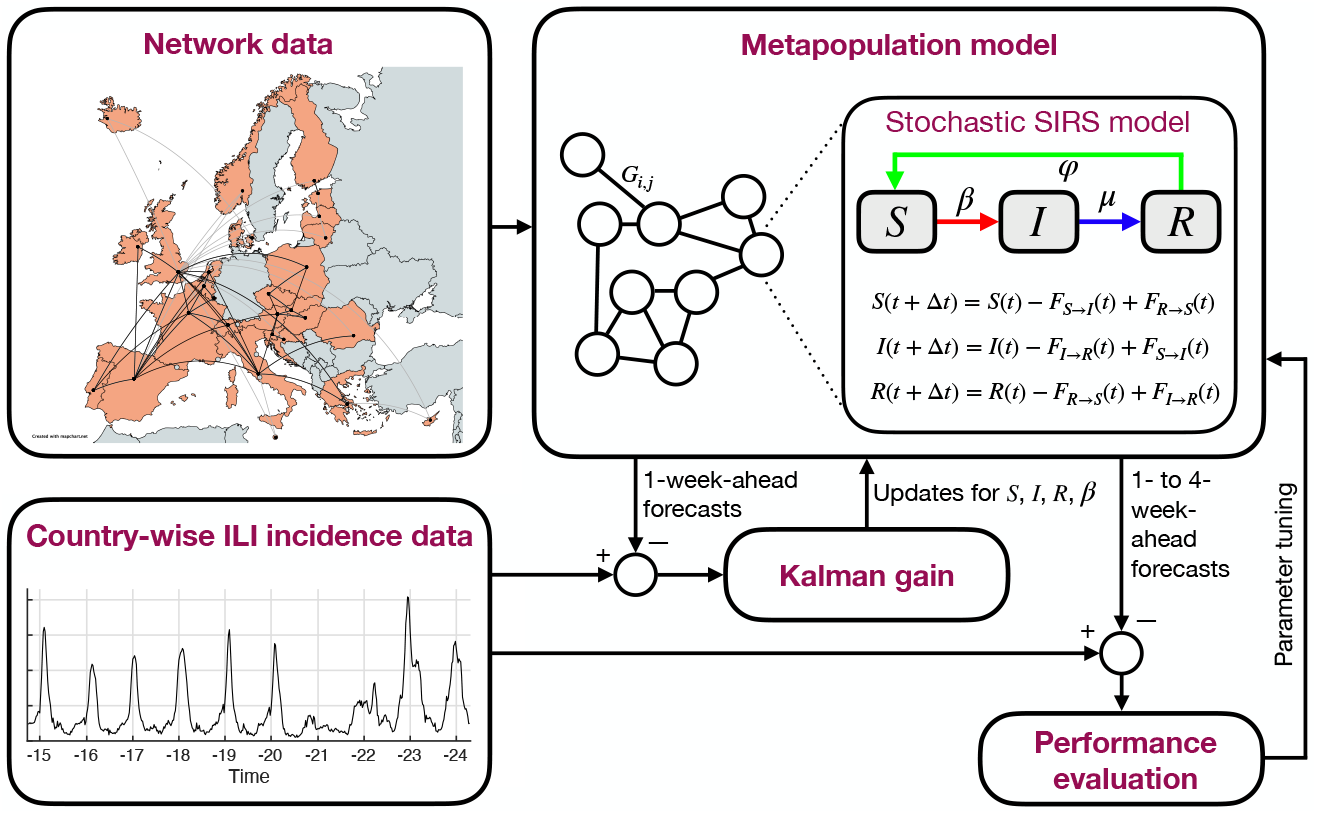
Model scheme. The network data panel shows daily fluxes above 10,000 with black lines, and the two strongest connections for each country with gray lines. Parameter tuning is described in Supplement S4.

### 2.3 Model development

After constructing the mobility network, we developed a dynamical model to simulate the spread of ILI across 28 European countries. The model is based on a stochastic discrete-time Susceptible-Infectious-Removed-Susceptible (SIRS) framework, which describes the time evolution of the disease spread in homogeneous and well-mixed populations [28, 29, 30, 31], extended to incorporate network effects. This captures both local epidemic dynamics within each country and the influence of cross-border interactions. The model, schematically represented in Figure3, is described in detail in Supplement S2.

#### Network-based SIRS dynamics

The model represents each country as a node in the network, with epidemic dynamics described by SIRS equations. Every country has a specific epidemic evolution that is affected by the epidemic states of other countries. The number of new infections in country *i*, that is, the flux from the susceptible population *S*_*i*_(*t*) to the infectious population *I*_*i*_(*t*) at time *t*, is given by

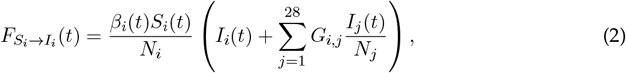

where *β*_*i*_(*t*) is the transmission rate within country *i, G*_*ij*_ is the flux of individuals from country *j* to country *i*, and *N*_*j*_ is the population of country *j*. The other fluxes are as in the standard SIRS model, 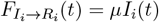 and 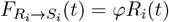.

#### Mean-field approximation

To explore the trade-off between data availability and model complexity, we implemented a simpler “mean-field” version of the model, where the detailed network matrix *G* is replaced with

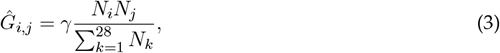

where γ is a tuning parameter. When this is substituted into Eq. (2), the outside effect reduces to the average incidence over all countries. This approach is less granular, but requires fewer data and is suitable for scenarios with incomplete mobility information.

#### Individual models

The performances of the network-based and mean-field models were compared against country-specific isolated models that do not consider cross-border interactions, that is, *G* is set to zero in Eq. (2).

### 2.4 Data integration by the Extended Kalman filter

To estimate epidemic parameters and make accurate predictions, the network-based SIRS model was implemented in a stochastic framework and coupled with an Extended Kalman Filter (EKF) [32], described in Supplement S3. The EKF integrates real-world observations with the model dynamics, providing robust state estimation and allowing for dynamic adjustments as new data become available.

#### State estimation

The EKF tracks the evolution of the state variables (susceptible, *S*; infectious, *I*; removed, *R*) and of transmission rate (β) at each node in the network. For each country, the EKF optimally combines noisy observational data with the model’s predictions to estimate the most likely epidemic state. This recursive process involves a prediction step using the model, and then an update of the state vector based on discrepancies between predictions and observations, ensuring that the model evolution is close to real-world dynamics.

#### Stochastic dynamics

To account for uncertainties in epidemic progression, the model incorporates stochastic noise into the transitions between compartments. For example, following the derivation of the Langevin equation in [33], infections are modeled as independent stochastic events, leading to a binomial distribution for the number of new infections, reflecting the inherent variability in disease spread. Similarly, case detections are modeled as stochastic events giving rise to a measurement noise model. The EKF framework explicitly accounts for these uncertainties through covariance matrices for both state noise and measurement noise. These matrices, calibrated to reflect the variability in the underlying data, are updated dynamically to capture changing epidemic conditions.

#### Dynamic transmission rates

The transmission parameters *β*_*i*_(*t*) in (2) are modeled as time-varying variables influenced by both local and network-level factors. Changes in *β*_*i*_(*t*) can reflect shifts in population behaviour, interventions, or seasonal effects. The dynamics of *β*_*i*_(*t*) are governed by

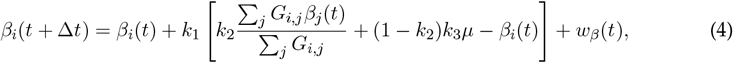

where the term inside the brackets acts as a nudge towards a reference value that depends on the weighted average of *β*_*j*_ in the neighbours of country *i*, while *k*_1_, *k*_2_, and *k*_3_ are tuning parameters, *µ* is the rate parameter for the fluxes *I*_*i*_ *→R*_*i*_, and *w β* (*t*) is Gaussian noise. In isolated models, *k*_2_ is zero. Including network effects in the -parameter dynamics differs significantly from previous models [19, 20, 21] as it allows regional waves to influence neighbouring countries, thus enhancing the model’s ability to anticipate epidemic patterns.

#### Probabilistic forecasting

The stochastic model can generate ensembles of future scenarios to generate probabilistic forecasts. We generate probabilistic forecasts by simulating 1,000 epidemic trajectories forward in time, starting from an initial state sampled from a multivariate normal distribution defined by the current state estimate as the mean and its error covariance matrix obtained from the EKF. Both stochastic dynamics and measurement noise are incorporated. This approach enables the quantification of uncertainties in the model’s predictions. An additional noise calibration step is required for the probabilistic forecasts, as described in Supplement S6.

#### Model implementation and validation

The SIRS model was implemented with a one-day time step (Δ*t* = 1*/*7, since we use a week as a time unit). Observational data, provided as weekly aggregated case numbers, were incorporated into the model through detection probabilities (*c*_*i*_(*t*)) that link reported cases to the underlying epidemic state by *y*_*i*_(*t*) = *c*_*i*_(*t*)*I*_*i*_(*t*) for *t ∈* ℕ, thereby adjusting for under-reporting (see Supplement S4 for details). These probabilities are estimated adaptively. In total, 11 parameters are used as tuning parameters (Supplementary Table S1), fitted with simulated annealing (Supplementary Table S2). Data from the first four epidemic seasons (2014–2018) are used for parameter fitting, and the remaining six seasons (2018–2024) are used for evaluations, with a special focus on critical periods such as wave onsets and epidemic peaks.

## 3 Results

### 3.1 Including network effects improves forecasting performance

#### Evaluation metrics and setup

Similarly to earlier works such as Influcast [6], we evaluated forecasting performance using 4-week-ahead predictions for influenza-like illnesses (ILI) across Europe. Forecasting performance was assessed from the adjusted average error between model predictions and actual data. Performance gain was quantified as the percentual reduction in error compared to a reference forecast, which uses the most recent observed incidence as the future prediction (details in Supplement S5). This evaluation was conducted over ten years of data, with separate analyses for training and test datasets, and for critical periods, namely wave onsets and epidemic peaks (Figure 2).

#### Model comparisons

Figure 4 illustrates the performance of the network model, mean-field model, and isolated models for 4-week-ahead forecasts. The network model consistently outperformed isolated models across all periods. Around epidemic peaks, the network model achieved up to a 25% improvement in forecasting performance. Wave onset remains difficult to forecast, but in these periods, the network model clearly outperformed the mean-field and isolated models.

**Figure 4:**
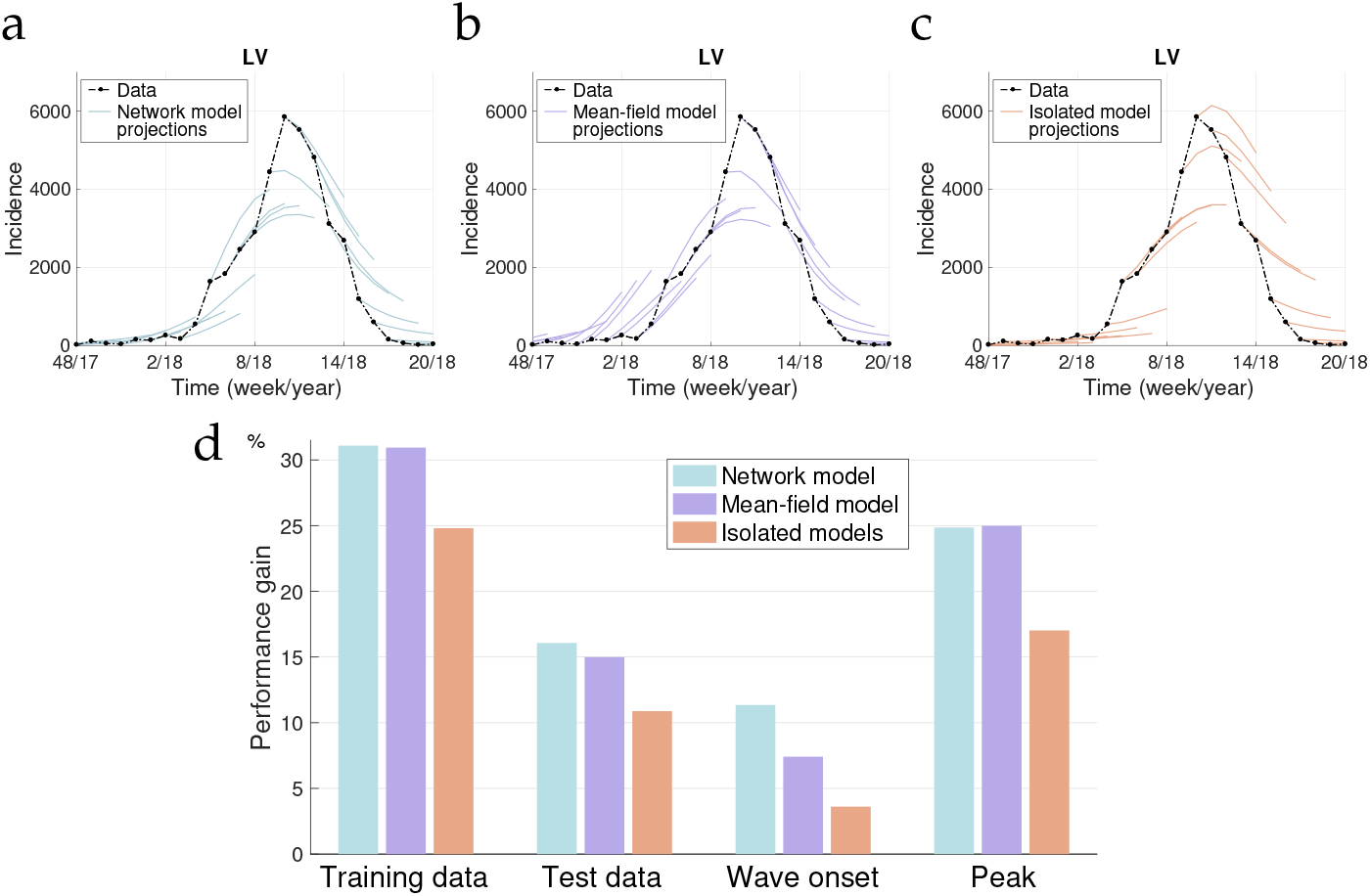
Comparison of 4-week-ahead forecasts across different models for the 2017–2018 epidemic season in Latvia. **a, b, c:** Forecasts generated by the network model, mean-field model, and isolated model, respectively. The network model accurately captures the wave onset, outperforming the other models, which either underestimate or misalign the timing of the initial wave. In this epidemic season, the epidemic wave reached the Baltic states later than elsewhere in Europe (*cf*. Figure 2), which explains the differences in the forecasts. **d:** Performance gain of each model compared to the reference forecast, across training and test data, as well as during wave onset and peak periods. These results highlight the largely superior predictive accuracy of the network model with respect to isolated models, particularly during critical epidemic phases.

#### Regional and temporal analysis

Performance gains varied by region and epidemic season (Figure 5). The network model provided the highest gains in countries with strong cross-border interactions, such as Austria and Slovakia. Predictions were more accurate during prepandemic seasons (2018–2019) compared to COVID-affected years, where reduced mobility and reporting inconsistencies impacted performance: since the mobility network is assumed to be static, it was less accurate when travel restrictions were in place. Also, the adaptive estimation scheme for the detection parameters *c*_*i*_(*t*) may be inadequate in extreme situations. Future improvements could incorporate a dynamic network evolution to reflect real-time changes in mobility patterns.

**Figure 5:**
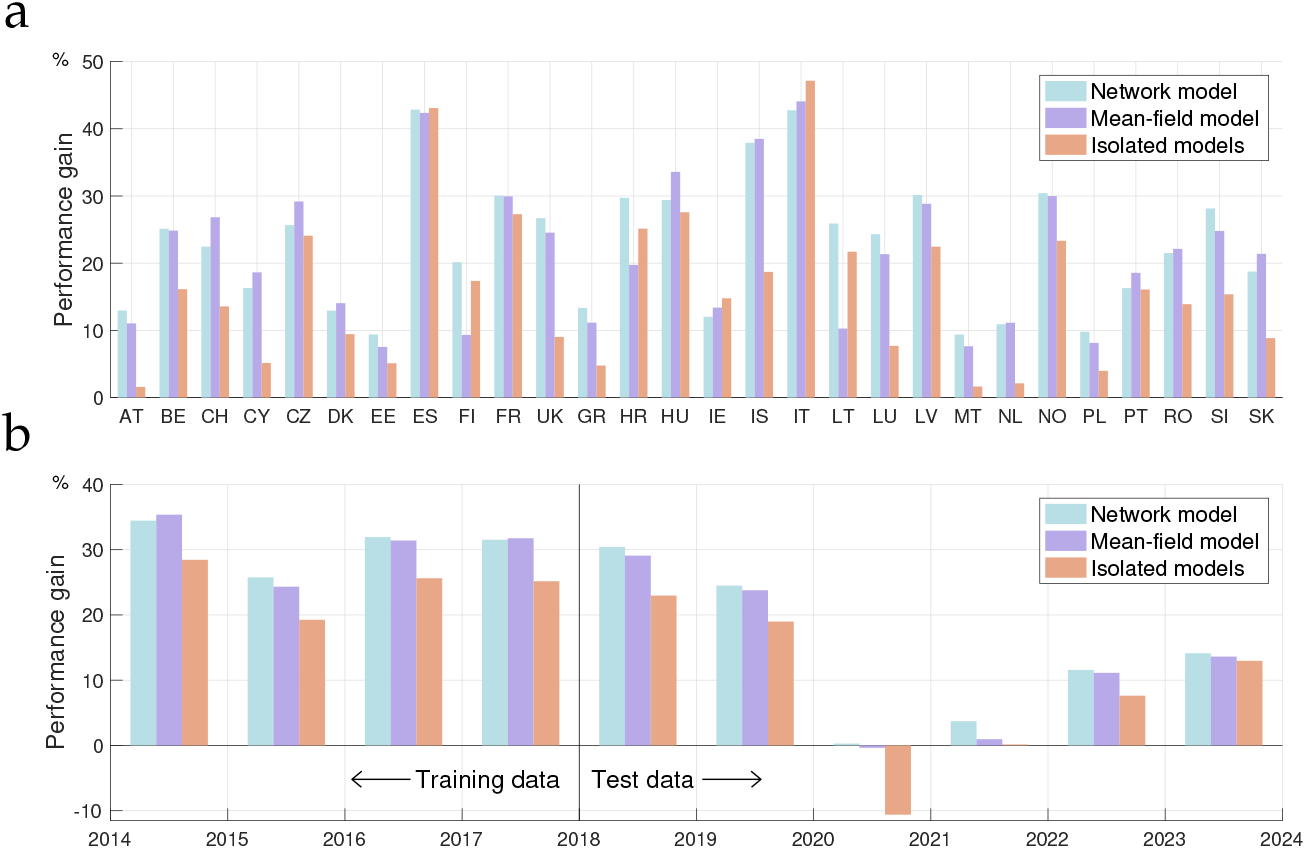
**a:** Performance gain for each country over the entire ten-year period. **b:** Season-by-season performance gains.

We also fitted model parameters using all ten years of data, to check whether providing longer time series improved model predictions. The performance did not change considerably, and the same drop in the performance gain for test data period compared to training data period could still be observed. This confirms that the pandemic was the main cause of this performance drop, rather than potential overfitting. A similar performance was obtained even when only the first two years of data were used for parameter fitting. This is indicative of the parsimony of our model, and suggests that some country-specific parameters might be fitted based on forecasting performance without risk of overfitting. Details of these experiments are in Supplement S7 and Supplementary Figure S3.

#### 3.2 Including network effects improves forecasting with missing data

It is crucial for an epidemic model to provide robust forecasts, even with missing data. Our network-based models leverage information from interconnected regions to impute missing data and provide accurate predictions, even when substantial portions of the dataset are unavailable.

#### Experiment setup

We simulated missing data scenarios by randomly deleting 20%, 40%, 60%, and 80% of the ILI data across all countries. Data that followed a period of at least three weeks of missing data were “protected” from deleting. Each scenario was repeated 20 times to account for variability. Forecasting performance was evaluated as described above, throughout the ten years considered. In addition, we measured how well the deleted data could be reconstructed using all available data until the reconstruction time. To enable direct comparison, the same error metric is used as for the forecasts.

#### Performance analysis

As shown in Figure 6a, the forecasting performance of the network-based model is robust to increasing percentages of missing data: prediction errors are rising only modestly. For example, with 40% data missing, the error increased by just 11.6%, compared to 15.1% for isolated models. The network model thus shows its ability to utilize inter-country dynamics to compensate for missing information. The reconstructed data closely aligned with true infection trends. Notably, reconstruction errors remained significantly lower than forecasting errors.

**Figure 6:**
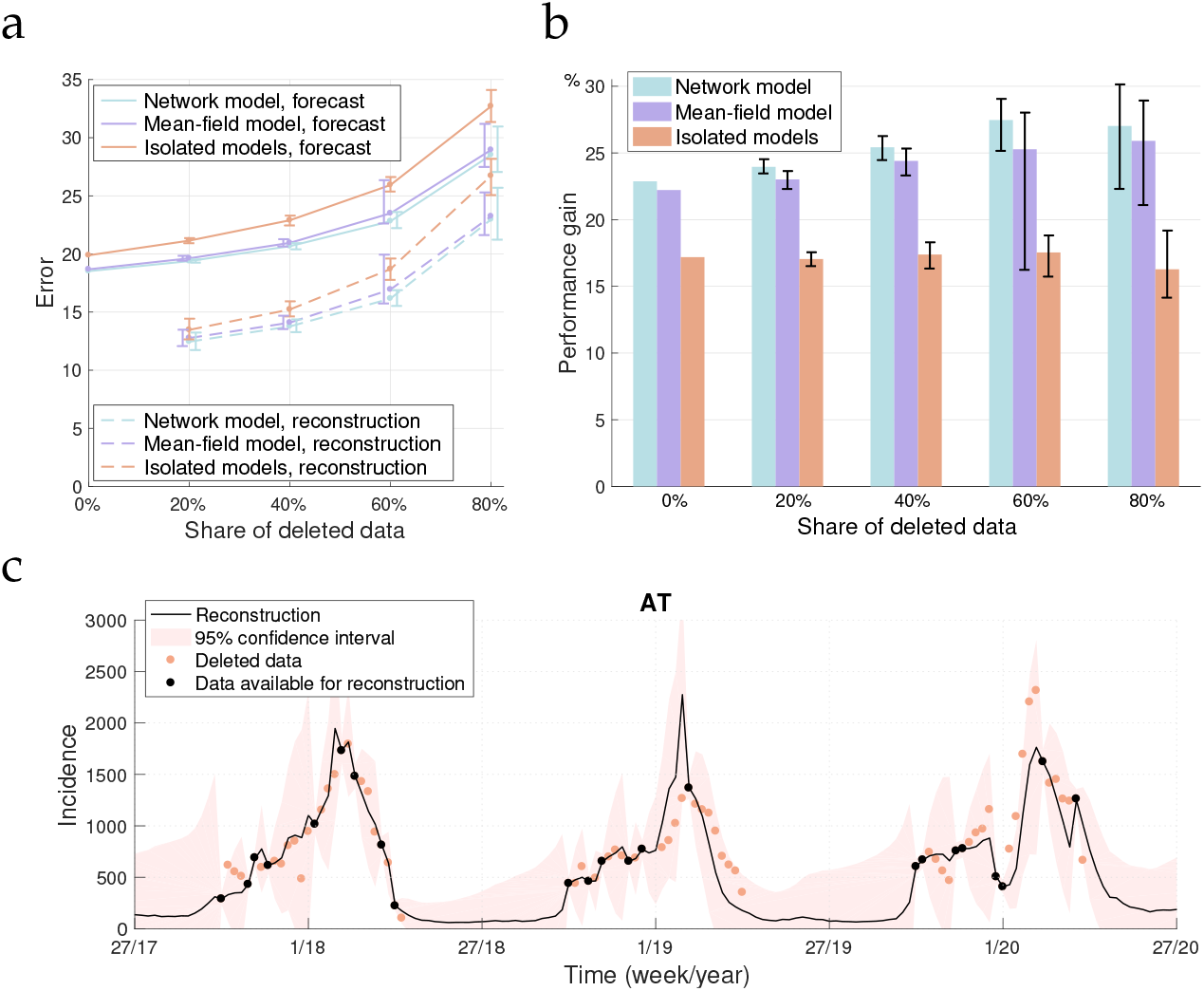
Performance of the models with different percentages of missing data. Confidence intervals show the 90th percentiles over different randomized evaluations. **a:** Total error for different percentages of deleted data; the network model has a consistently better performance, the error remaining relatively low even with 80% missing data. **b:** Performance gain with missing data compared to the reference forecast. **c:** Example of data reconstruction for Austria over three epidemic seasons with 60% of the data deleted. The network model effectively reconstructs the missing data while maintaining confidence intervals. Observe the increasing width of the confidence interval in periods when data are missing.

#### Benefits of network integration

The performance gain of the network model over the reference forecast increased with higher deletion levels, rising from 23% to 27.5% as data availability decreased from 100% to 40% (Figure 6b), while the performance gain of the isolated models remained roughly constant at around 17%. Figure 6c shows an example for three epidemic seasons in Austria with 60% of the data deleted.

Our findings demonstrate the value of integrating information from neighbouring countries, particularly in scenarios with incomplete datasets, and hence the utility of network-based models for improving forecasts and imputing missing data, which is particularly relevant for epidemic monitoring in regions with uneven surveillance or reporting practices.

### 3.3 Probabilistic forecasts

Uncertainty quantification provided by the Extended Kalman Filter allows the network-based model to provide valuable insight into the potential future trajectory of epidemics, accounting for inherent uncertainties and generating confidence intervals and probabilistic scenarios to guide decision-making.

Figure 7 illustrates examples of probabilistic forecasts for France and Switzerland during the 2023–2024 epidemic season. The model provides point estimates and different confidence intervals, capturing the uncertainty in the disease progression. The forecasts demonstrate the ability of the network model to anticipate epidemic trends, including wave peaks and durations, while offering actionable ranges for public health planning.

**Figure 7:**
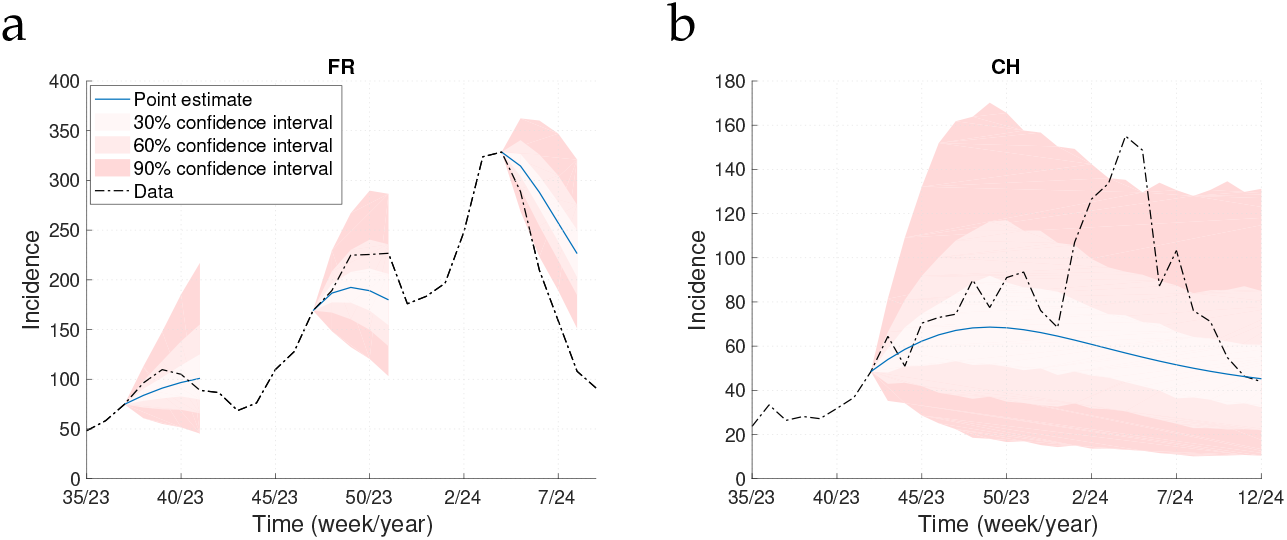
Probabilistic forecasts for the 2023–2024 epidemic season in France and Switzerland. **a:** 4-week-ahead probabilistic forecasts for France. **b:** Long-term probabilistic scenario simulation for Switzerland.

### 3.4 Insights into model mechanisms and interpretability

One of the strengths of the network-based model lies in its mechanistic design, which allows for a detailed inspection of the factors driving its performance. To better understand these dynamics, we conducted additional experiments, described in detail in Supplement S7.

Firstly, we investigated the role of population sizes by scaling the network matrix *G* row-wise. The results indicated that population size does not significantly influence forecasting performance, suggesting that the model’s accuracy stems primarily from the structural and dynamic properties of the network itself.

Secondly, the inclusion of network effects in the dynamics of the transmission parameter *β* was found to play a crucial role in enhancing forecasting capabilities. When this effect was removed by setting *k*_2_ =0 in (4), the model’s performance dropped markedly.

Finally, we tested a hybrid model where the mean-field network informed only the *β* - parameter dynamics, while the *S →I* transitions were governed by isolated models. The performance of this model was mostly comparable to the full network model. Interestingly, the detailed network model outperformed all alternatives during wave onset periods, highlighting its strength in capturing the early dynamics of epidemic waves.

## 4 Discussion

### Summary

We demonstrated the advantages of a networked metapopulation SIRS model, integrated with an Extended Kalman Filter (EKF), for forecasting influenza-like illnesses (ILI) across Europe. The model leverages empirical data on mobility and disease incidence to incorporate both local dynamics and cross-border interactions and thus improve forecasting accuracy. Comparative analyses show that the network-based model outperforms isolated and mean-field models, particularly during critical periods such as wave onsets and epidemic peaks. Additionally, the model is robust to missing data and provides reliable forecasts even with substantial data unavailability.

### Conclusions and future impact

Our findings highlight that integrating mobility data into network-based epidemic models improves accuracy and interpretability. In a probabilistic forecasting framework, the EKF enables the model to adapt dynamically to new data, providing real-time forecasts with quantified uncertainty that is critical for proactive responses. Our proposed framework is highly relevant for epidemic monitoring and regional and global public health decision-making, with several applications in forecasting. By providing a spectrum of plausible epidemic trajectories, it supports scenario analysis and planning, including: 1) Early warnings: identifying potential wave onsets and peak periods with quantified uncertainty; 2) Resource allocation: informing healthcare capacity planning by projecting worst-case and best-case scenarios; 3) Scenario modelling: evaluating the impact of interventions, such as travel restrictions or vaccination campaigns, by simulating alternative futures.

The demonstrated benefits of network-based approaches are expected to influence the design of future epidemic forecasting systems, particularly in interconnected regions. Our results encourage the adoption of similar frameworks for diseases such as COVID-19, dengue, and seasonal influenza. The reconstructed European mobility network can also serve as a foundation for further studies, fostering collaboration and data sharing among public health agencies.

### Limitations and future work

Static network structures limit the accuracy of predictions during periods of rapid mobility changes, such as those caused by pandemics or natural disasters. Incorporating dynamic network adjustments, informed by real-time mobility data, could enhance model adaptability and performance. Our pipeline also implicitly assumes that data collection is consistent between epidemic seasons. The adaptive scheme for estimating the detection parameters *c*_*i*_(*t*) may not be able to cope with drastic changes that are sometimes observed in the data. Any knowledge on changes can be manually incorporated to improve performance.

Moreover, the model currently assumes homogeneous mixing within populations, which may oversimplify complex interactions, particularly in urban areas. Future work could integrate finer-grained data, such as inter-city mobility patterns, or other complementary data such as holiday schedules like in [30], or humidity data like in [4, 18, 21, 30]. Additionally, the absence of certain countries from the network, due to data unavailability, may introduce regional biases. Expanding public databases and standardizing data collection practices across Europe would address this issue.

Lastly, while the model performs well for ILI, its applicability to other diseases with different transmission dynamics, such as vector-borne or chronic diseases, remains to be explored. Adjusting the framework to accommodate diverse epidemiological characteristics would broaden its impact.

## Data Availability

Used data are available at https://gitlab.com/uniluxembourg/lcsb/systems-control/epinetekf

## Data and code availability

The code is available at gitlab.com/uniluxembourg/lcsb/systems-control/epinetekf together with a user guide and the data used for the results in this article.

## Acknowledgements

A.A. was supported by the Luxembourg Government through the CoVaLux programme (16954531). D.P. and G.G. are funded by the European Union through the ERC INSPIRE grant (project number 101076926). Views and opinions expressed are however those of the authors only and do not necessarily reflect those of the European Union or the European Research Council Executive Agency. Neither the European Union nor the European Research Council Executive Agency can be held responsible for them. The authors wish to express their gratitude to the team behind the RespiCast hub.

## Data sources

- **Flight data:** Eurostat, Air passenger transport between reporting countries, Link, doi: doi.org/10.2908/avia_paocc, [Accessed 2024-08-22].
- **Ferry traffic data:** Finnish Port Association, statistics, Link, [Accessed 2024-08-23].
- **Ferry traffic data:** Sea Passenger Statistics: All routes 2019, UK Government, Department for Transport, Link, [Accessed 2024-08-23].
- **Epidemic data:** European Centre for Disease Prevention and Control, European respiratory virus surveillance summary (ERVISS). Link, [Accessed 2024-09-23].
- **Epidemic data:** World Health Organization, Global influenza programme, FluID, Link, [Accessed 2024-09-23].
- **Population density data:** Eurostat, Population density by NUTS 3 region, Link, [Accessed 2024-08-28].

## Notes

### Competing Interest Statement

The authors have declared no competing interest.

