## Supplementary material for "Networked SIRS model with Kalman filter state estimation for epidemic monitoring in Europe"

**Note:** Citations in the Supplementary Material refer to the list of references in the Main Text. There are no references appearing only in the Supplementary Material.

### S1 Network generation

Data for plane travels in 2023 were obtained from the Eurostat Data Browser, "Air passenger transport between reporting countries"; only for UK, we used data from 2019 as the most recent available data before the COVID-19 pandemic. The flight data are given quarterly, and we averaged them over the whole year. Some missing value imputation is done based on data from different quarters between the countries in question, accounting for overall quarterly differences. Ferry traffic data were gathered from the statistics of the Finnish Port Association for Finland[1] and from the UK Government, Department for Transport *Sea Passenger Statistics: All Routes 2019* report. Air and maritime traffic were disrupted by the COVID-19 pandemic, and recent data may diverge from actual pre-pandemic traffic data. We were able to capture earlier trends by using averages, as well as by tuning extra parameters $\gamma_{1,2}$ in our model (see below). Future studies may embed dynamical data flows, when available for the whole network.

Data for cross-border worker flows were obtained from the Directorate-General for Employment, Social Affairs and Inclusion of the European Commission [26] for the EU and EFTA areas, and from the Economic and Social Research Institute [27] for the UK-Ireland border. The worker flows between non-neighbouring regions were omitted, since these flows should in most cases appear in the travel statistics. It was assumed that cross-border workers actually cross borders on 60% of days (thus excluding weekends, bank holidays and average leave periods). Cross-border workers were counted in the network as flows in both directions, between their country of residence and the country of work, as they were assumed to commute daily and thus be able to carry an infection in either direction.

Finally, the neighbour graph lumps together all other travels across the land borders of different countries in the absence of available data. These links were constructed based on population densities in border regions, together with information about the border length. The latter was informed by the NUTS 3 (Nomenclature of Territorial Units for Statistics) regional classification[2]. This procedure, further scaled via parameter fitting as described in Main Text, provides estimates for unregistered movements. Having better curated datasets may further improve this estimation in future applications. Further details about the construction of the neighbour graph were provided below.

#### S1.1 Construction of neighbour graph

The neighbour graph was not directly established from public databases, but is imputed from proxy data. In particular, for each NUTS 3 region that shares a border with another country, we measured the length of the border segment between the region and the neighbouring country, and weighted that with the population density of the region (obtained from demographic databases). Weighted segments were then added up for the whole border between two countries, including the border regions from both countries so as to have a symmetric network matrix. The numbers obtained in this way are considered as dimensionless units and later scaled by parameter fitting. An example for this procedure for the Czechia–Slovakia border is shown in Supplementary Figure S1.

This procedure provides a simple estimation of unregistered movements, but it has some shortcomings. For example, it looks at only one side of the border at a time. In reality, there is likely more traffic across the border if both sides of the border are densely populated. For

[1] https://www.finnishports.fi/eng/statistics/trade-information/port-of-helsinki-top-in-europe/ [Accessed 2024-08-23]

[2] https://doi.org/10.2785/714519 [Accessed 2024-08-28]

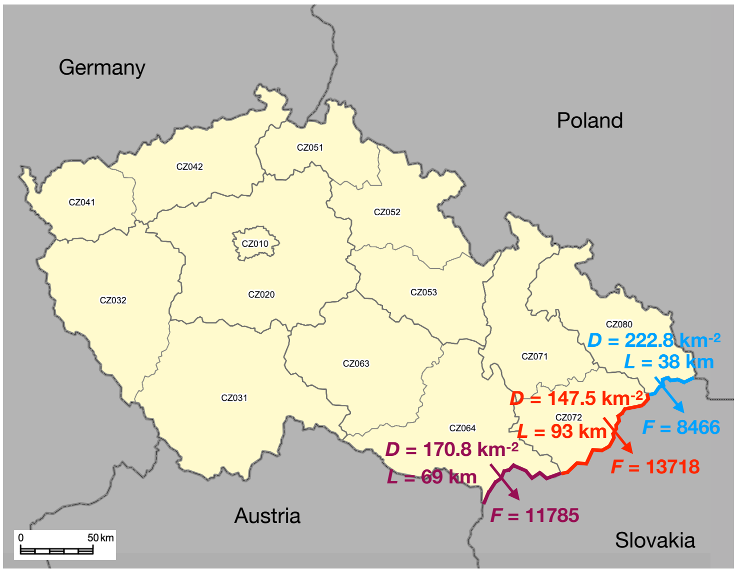


Supplementary Figure S1: NUTS 3 classification of regions for Czechia, and an illustration on the construction of the neighbour graph for the Czechia–Slovakia border. The flux across the border ($F$) is obtained as the product of the region's population density ($D$) and the length of the border segment between the region and the neighbouring country ($L$). The map is from NUTS (https://ec.europa.eu/eurostat/web/nuts ▷ Maps), with our own additions.

example, Vienna and Bratislava, capitals of Austria and Slovakia, are located very close to each other. While our procedure is of course predicting fairly high traffic across the border, it might still be under-estimated. Another simplification is caused by not taking into account geographical specificities. For example, the borders between France and Italy and France and Spain are on mountainous regions. Even though both sides of these borders are densely populated, the traffic across these borders might be over-estimated by our procedure. Having better curated datasets may further improve this estimation in future applications.

Some outlier flows appear by this procedure. These outliers arise due to densely populated regions, which have a narrow or protruding shape and share a relatively long border with a neighbouring country. In addition, some cities constitute their own regions in the NUTS 3 classification, and if these cities lie at the border, they may cause outliers as well. These outliers are dealt with either by joining the corresponding regions with (larger) neighbouring regions or, in few cases, by manually modifying the border length.

For example, on the Italian–Slovenian border, the Trieste province (NUTS3 code ITH44) is very densely populated and narrow, being only a couple of kilometers wide. Its neighbouring province is Gorizia (ITH43), whose size is relatively small as well. Flows between these regions and Slovenia are thus outliers in our method. Hence, both Trieste province and Gorizia province were joined with the larger Udine province (ITH42) to obtain reasonable flow estimates. Similarly, the Geneve canton (CH013) on Switzerland's border with France was joined with the neighbouring Vaud canton (CH011), and the canton of Basel–Stadt (the city of Basel, CH031) was joined with the surrounding canton of Basel–Land (CH032). On the French–Belgian border, the densely populated and narrow department of Nord (FRE11) was joined with the neighbouring department Pas-de-Calais (FRE12), even though the latter does not share a border with Belgium.

The Zuid–Limburg region (around Maastricht) in the Netherlands forms a protrusion into

Belgium, and has a very high population density. To account for the protruding shape, the length of the border between the Zuid–Limburg region and Belgium was reduced by a third. The border length between the Torino province (ITC11) and France was reduced from 141 km to 111 km, excluding some excessive turns the border makes. Similarly, the border length between the Belgian arrondissement of Antwerp and the Netherlands was reduced from 51 km to 32 km, excluding some turns.

### S2 Model development

#### S2.1 SIRS model at each node of the network

We employ a discrete-time SIRS model to capture the time-scale of data sampling:

$$\begin{aligned} S(t+\Delta t) &= S(t) - F_{S\to I}(t) + F_{R\to S}(t) \\ I(t+\Delta t) &= I(t) + F_{S\to I}(t) - F_{I\to R}(t) \\ R(t+\Delta t) &= R(t) + F_{I\to R}(t) - F_{R\to S}(t). \end{aligned} \tag{1}$$

where $F_{X\to Y}(t)$ is a shorthand notation for the flow from compartment $X$ to compartment $Y$ at time $t$, with unit of one week (we will later omit the time-dependency). The model's time step is one day, $\Delta t = 1/7$. The model satisfies the conservation law

$$S(t+\Delta t) + I(t+\Delta t) + R(t+\Delta t) = S(t) + I(t) + R(t).$$

In the country-specific SIRS model, the fluxes between compartments are given by $F_{S\to I}(t) = \beta(t)S(t)I(t)/N$, $F_{I\to R}(t) = \mu I(t)$, and $F_{R\to S} = \phi R(t)$. In the metapopulation network model, epidemic dynamics in $M$ regions with populations $N_i$ ($i = 1\ldots M$) are each modeled by SIRS equations, but the regions are connected by effective flows $G_{i,j}$ of individuals from region $j$ to region $i$ per day. The flows are called "effective" since we will estimate global parameters multiplying the network matrix based on forecasting performance. In the network model, the number of new infections per time unit in country $i$ is given by

$$F_{S_i\to I_i}(t) = \frac{\beta_i(t)S_i(t)}{N_i}\left(I_i(t) + \sum_{j=1}^{M} G_{i,j}\frac{I_j(t)}{N_j}\right), \tag{2}$$

that is, it depends on the infectious population within the country $i$ itself, but also other countries: $I_j(t)/N_j$ is the fraction of infected individuals in country $j$; hence assuming perfect mixing of populations, the flux of infected individuals from country $j$ to country $i$ is $G_{i,j}I_j(t)/N_j$. We assume that these flows are symmetric: $G_{i,j} = G_{j,i}$. This assumption is a slight simplification of a more general conservation law stating that the population of each country remains constant, which would require $\sum_i G_{j,i} = \sum_i G_{i,j}$ for all $j$. We do not differentiate between people traveling to another region and becoming infected there, and infectious people traveling and infecting others at the destination region.

For the mean-field network model, instead, we substitute $G$ defined in Eq. (3) of Main Text into (2) to get

$$\hat{F}_{S_i\to I_i}(t) = \beta_i(t)S_i(t)\left(\frac{I_i(t)}{N_i} + \gamma\frac{\sum_{j=1}^{M} I_j(t)}{\sum_{j=1}^{M} N_j}\right) \tag{3}$$

where the second term inside the parentheses does not depend on the target country $i$, but is the average incidence over all included countries, modulated by the parameter $\gamma$. This model ignores geographical and other detailed information on the traffic between countries.

#### S2.2 Stochastic model

Towards an implementation of the EKF, a stochastic version of the networked SIRS model (2) is obtained based the method described in [24]. We follow the derivation of a Langevin equation [33], that is, we pose that each susceptible person has a probability (in the non-networked version, for simplicity) $\beta(t)I(t)/N$ to become infected at time $t$. The number of new infections on a day at time $t$ is then binomially distributed with mean $\frac{\beta(t)I(t)}{N}S(t)$ and variance $\frac{\beta(t)I(t)}{N}\left(1-\frac{\beta(t)I(t)}{N}\right)S(t)$. Assuming $\frac{\beta(t)I(t)}{N} \ll 1$, the term with $\left(\frac{\beta(t)I(t)}{N}\right)^2$ is omitted from the variance. Further assuming $\frac{\beta(t)I(t)}{N}S(t)$ is large enough, the binomial distribution can be well approximated by the normal distribution with the same mean and variance.

As a result, each transition between compartments is accompanied by a stochastic white noise process modulated by a multiplicative state-dependent term, which scales as the square root of the transition rate. To account for the fact that the model is based on the assumption of well-mixed homogeneous populations, the variance terms are multiplied by a tuning factor $\kappa_1$. In addition, a second term $\kappa_2 N_i^2$ is added to the variance of the transition from $S$ to $I$, that does not depend on the epidemic state. This can be interpreted as import of cases from countries that are not included in the network; a second reason behind its introduction is technical: during summer periods, many countries stop the ILI monitoring and reporting, hence the model tends to a state where there are no new cases emerging. Then, also the uncertainty on the epidemic state eventually converges to zero. When reporting is restarted, the Kalman filter can mainly make updates on the $\beta$-parameter due to its unfounded certainty on the model variables. This leads to poor projections in the beginning of a new epidemic season. The term $\kappa_2 N_i^2$ is introduced to mitigate this problem by providing inertia to uncertainties at the beginning of a new season. The final stochastic networked SIRS model is:

$$\begin{aligned} S(t+\Delta t) &= S(t) - F_{S\to I} + F_{R\to S} - \sqrt{\kappa_1 F_{S\to I}}w_1(t) + \sqrt{\kappa_1 F_{R\to S}}w_3(t) - \sqrt{\kappa_2}Nw_4(t) \\ I(t+\Delta t) &= I(t) + F_{S\to I} - F_{I\to R} + \sqrt{\kappa_1 F_{S\to I}}w_1(t) - \sqrt{\kappa_1 F_{I\to R}}w_2(t) + \sqrt{\kappa_2}Nw_4(t) \\ R(t+\Delta t) &= R(t) + F_{I\to R} - F_{R\to S} + \sqrt{\kappa_1 F_{I\to R}}w_2(t) - \sqrt{\kappa_1 F_{R\to S}}w_3(t). \end{aligned} \tag{4}$$

where $w_k(t)$ are mutually independent Gaussian white noise processes with unit variance.

#### S2.3 Dynamics of transmission parameter

Due to changes in behaviours, interventions and other factors, the transmission parameters $\beta_i(t)$ may vary in time. The stochastic SIRS model is thus complemented with a dynamical equation for the transmission parameter $\beta(t)$. In contrast to the model in [24], we include deterministic dynamics for the $\beta$-parameters. The reason for the inclusion is twofold. Firstly, due to missing data in many countries during the summer periods, the Kalman filter does not make significant updates to the $\beta_i$ parameters during these periods. However, it is reasonable to assume that, before seasonal re-emergence of ILI diseases, the effective transmission approaches similar values each year, without retaining the value reached at the end of the previous wave. The deterministic dynamics for $\beta_i$ first takes into account this seasonality. Secondly, by allowing the country-specific $\beta_i$ to be influenced by the parameters of the neighbours in the network, the parameter estimates become more stable. As seen in the results (Supplementary Figure S3), this feature also helps to anticipate a regional wave when that has been originated elsewhere.

The stochastic model used for $\beta_i(t)$ is

$$\beta_i(t+\Delta t) = \beta_i(t) + k_1\left(k_2\frac{\sum_j G_{i,j}\beta_j(t)}{\sum_j G_{i,j}} + (1-k_2)k_3\mu - \beta_i(t)\right) + w_5(t), \tag{5}$$

where $w_5$ is white noise with variance $\sigma_\beta$, and $k_1$, $k_2$, and $k_3$ are tuning parameters. The first term inside the parentheses is the weighted average of the $\beta_j$-parameters of the network neighbours, and the second term $k_3\mu$ is a grounding value, towards which the $\beta$-parameters tend to in the absence of data. As a consequence, if there were no data for any of the countries, the system would eventually converge to an endemic steady state, which is (for the isolated models) $S = N/k_3$, $I = \frac{\phi(k_3-1)}{(\phi+\mu)k_3}N$, and $R = \frac{\mu(k_3-1)}{(\phi+\mu)k_3}N$.

#### S3 Extended Kalman filter implementation

An Extended Kalman filter algorithm [32] requires an underlying dynamical model (in our case, the stochastic networked SIRS model), its output, covariance matrices for state noise and measurement noise, and measurement data. The EKF allows to estimate the state of the SIRS system, based on available data, by evaluating the optimal value of a set of variables of interest, to reproduce the data while accounting for uncertainties. To apply the Extended Kalman filter into the networked system, the state of the system for each country is represented with a 3-dimensional state vector, consisting of $S_i(t)$, $I_i(t)$, and $\beta_i(t)$. The $R_i(t)$ compartments are not included, since they can be recovered using conservation of mass, $R_i(t) = N_i - S_i(t) - I_i(t)$.

The 3-dimensional state vectors for all 28 countries are concatenated into one 84-dimensional state vector $x$, for which we have a stochastic model

$$x(t + \Delta t) = f(x(t)) + W(t).$$

The function $f : \mathbb{R}^{84} \to \mathbb{R}^{84}$ is defined block-wise $f(x) = [f_1(x), ..., f_{28}(x)]^\top$ country by country through equations (1), (2), and (5). The state noise vector $W(t)$ includes the contributions of all noise terms in the stochastic model (4), and from the stochastic model we can obtain its covariance matrix $Q$ as a block diagonal matrix whose $i^{\text{th}}$ block is given by

$$Q_i = \begin{bmatrix} \kappa_1(F_{S_i\to I_i} + F_{R_i\to S_i}) + \kappa_2 N_i^2 & -\kappa_1 F_{S_i\to I_i} - \kappa_2 N_i^2 & 0 \\ -\kappa_1 F_{S_i\to I_i} - \kappa_2 N_i^2 & \kappa_1(F_{S_i\to I_i} + F_{I_i\to R_i}) + \kappa_2 N_i^2 & 0 \\ 0 & 0 & \sigma_\beta \end{bmatrix}.$$

The observed case numbers are also assumed to be affected by uncertainties. We assume that every infectious person has a probability $c_i(t)$ to be detected at week $t$ and the detections are independent. Then, the number of detected cases is binomially distributed with mean $c_i(t)I_i(t)$ and variance $(1 - c_i(t))c_i(t)I_i(t)$. The model-predicted number of detected cases for country $i$ is therefore given by

$$\hat{y}_i(t) = c_i(t)I_i(t)$$

and the full observation vector $\hat{y}(t) = [\hat{y}_1(t), ..., \hat{y}_{28}(t)]^\top$ is obtained as $\hat{y}(t) = C(t)x(t)$ where $C(t)$ is a $28 \times 84$ matrix whose element $(i, 3(i-1)+2)$ is $c_i(t)$ with zeros elsewhere. The measurement noise covariance matrix $U$ for the full observation vector is diagonal with elements

$$U_i(t) = \big(\rho_1(1 - c_i(t))y_i(t) + \rho_2 N_i\big)u_i \tag{6}$$

where the first term corresponds to the variance of the binomial distribution (using the observed number $y_i(t)$ as a proxy for $c_i(t)I_i(t)$). As we did with the stochasticity of new infections, here we also introduce a second, state-independent noise term whose variance is proportional to the population size $N_i$. $\rho_1$ and $\rho_2$ are tuning parameters for the different noise terms, and $u_i$ is a country-specific coefficient measuring the noise level in the data, defined by

$$u_i = \frac{1}{T_{\max}} \sum_{t=1}^{T_{\max}} \frac{(y_i(t) - \bar{y}_i(t))^2}{\bar{y}_i(t)}\ ,$$

where $\bar{y}_i(t)$ denotes the 5-week moving average of $y_i$, with the moving window centered at $t$.

The parameter $c_i(t)$ in the model output is also related to the ratio of detected and total cases, which is captured by $\frac{c_i(t)}{7\mu}$, and it is essential in modelling the wave amplitude. This parameter is time-varying due to an adaptive scheme used to estimate it, which is described in the following section.

As a small remark, note that the CoWWAn [24] model and the Respicast projections use a slightly more complex observation model with an additional "counter" variable for the daily or weekly new cases. However, we observed that the performance was just as good with a simpler model, where case numbers are simply obtained from the size of the $I$-compartment, which is what we currently implement.

If the covariance matrices $Q$ and $U$ and the initial uncertainty covariance $P_0$ are multiplied by a constant, then the error covariance $P_t$ at all times $t$ will be multiplied by the same constant, as will also the prediction error covariance $\tilde{P}$ and the measurement error covariance $S$ in Algorithm 1. In the Kalman gain matrix $\tilde{P}C(t)S^{-1}$, however, the constant will cancel out due to the inverse of $S$, so that the Kalman gain and, in turn, the state estimates $\hat{x}(t)$ remain unchanged by such scaling. In our model, we therefore keep $\sigma_\beta$ fixed to have some grounding value for the other covariance terms that are scaled by tuning parameters $\kappa_1$, $\kappa_2$, $\rho_1$, $\rho_2$. These, in turn, are optimized using noise-free projections, that do not change even if all covariances were scaled. In order to produce probabilistic forecasts, a final calibration step will be carried out to find a proper scaling factor for the covariances.

After every Kalman update, the model (without noise) is run 4 weeks forward in time to produce incidence projections for $\tau = 1, 2, 3, 4$ weeks ahead, denoted by $\hat{y}_i(t + \tau|t)$. That is, $\hat{y}_i(t + \tau|t)$ is a projection for week $t + \tau$ done using data up to week $t$.

```
Set P_0 ∈ ℝ^{84×84} and x̂(0);
while t ≤ T_max do
    set x̃ = x̂(t − 1);
    set P̃ = P_{t−1};
    for i=1,...,7 do
        Prediction error covariance: P̃ = J_f(x̃) P̃ J_f(x̃)^⊤ + Q(x̃);
        Prediction for time t − 1 + i/7: x̃ = f(x̃);
    end
    Measurement prediction error covariance: S = C(t) P̃ C(t)^⊤ + U(t);
    State update: x̂(t) = x̃ + P̃ C(t)^⊤ S^{−1} (y(t) − C(t) x̃);
    Error covariance update: P_t = P̃ − P̃ C(t)^⊤ S^{−1} C(t) P̃;
    if S_i ≥ 0.4 N_i for all i then
        t = t + 1;
    else
        Update c_i(t) upward;
        t = t − 12;
    end
end
```

**Algorithm 1:** The Extended Kalman filter for the networked SIRS model. $J_f(x)$ is the Jacobian of the function $f$, evaluated at $x$. The algorithm is standard, but the prediction step consists in solving the SIRS model one week forward in time between measurement times.

### S4 Parameters

#### S4.1 Adaptive estimation of $c_i(t)$

Key parameters in the method are the detection parameters $c_i(t)$. In SIR-type models, the initial exponential growth rate of a new wave and the herd immunity level are tightly connected. However, if only a fraction of the infections are detected, this has no effect on the observed initial exponential growth rate but it directly affects the amplitude of the upcoming (detected) wave. Without knowledge of this fraction, the link between initial growth rate and wave amplitude is broken, which precludes (model-based) projections going beyond short-term extrapolation.

Mostly, the wave sizes in a country do not vary greatly between consecutive years. Therefore an old parameter value obtained from previous years can be used as an initial parameter for a new epidemic wave. Sometimes, however, there are bigger changes between years, and therefore we have developed an adaptive scheme for updating these parameters, described in the paragraphs below.

**Initial values.** The initial values for the ratios are based on the observed epidemic dynamics for the first two epidemic waves in the data. The discounted case numbers are calculated as $Y_i(t; 1-\nu) = \sum_{\tau=0}^{t}(1-\nu)^{t-\tau} y_i(\tau)$ where $\nu = \frac{1}{1/\mu + 1/\phi}$ represents the average flow rate through the $I$ and $R$ compartments (that is, $1/\nu$ is the average time from infection to becoming susceptible again). Then

$$c_i(1) = \alpha_1 \frac{\max_{t \in [T_{0,i}, T_{0,i}+103]} Y_i(t; 1-\nu)}{N_i} \tag{7}$$

where $T_{0,i}$ denotes the first week when data are available for country $i$. Here $\alpha_1$ is a tuning parameter.

**Upward update.** If a too small value $c_i(t)$ is used for a country during an epidemic wave, the $S$-compartment will become exhausted. When this happens, more precisely, when $S_i(t) < 0.4N_i$ for some country, this is deemed unrealistic, and a parameter adaptation is triggered. The parameter $c_i(t)$ is retroactively increased, and the last 12 weeks are re-run with the higher parameter value. Further upward updates are not allowed during the re-run weeks.

The coefficient used for increasing $c_i(t)$ is determined as follows. The discounted true case numbers and 1-week ahead forecasts are calculated:

$$Y_i(t; 0.8) = \sum_{\tau=1}^{t} 0.8^{t-\tau} y_i(\tau), \qquad \hat{Y}_i(t; 0.8) = \sum_{\tau=1}^{t} 0.8^{t-\tau} \hat{y}_i(\tau).$$

The parameter $c_i(t)$ is then increased by multiplying it with the coefficient

$$\max\left\{1.1, \left(\frac{Y_i(t; 0.8)}{\hat{Y}_i(t; 0.8)}\right)^{\alpha_2}\right\} \tag{8}$$

where the exponent $\alpha_2$ is a tuning parameter used to link case numbers with the parameter $c_i(t)$. The parameter is increased retroactively in a piecewise linear fashion, that is, $c_i$ has a linearly increasing phase from $t-12$ to $t-6$ from the old value to the increased value. From week $t-5$ to $t+15$, the parameter remains at the new higher value. From week $t+16$ to $t+26$ the parameter linearly decreases to a value that is $c_i(t-12) + \alpha_3(c_i(t) - c_i(t-12))$, that is, $\alpha_3 \in (0, 1)$ denotes a share of the parameter update that is kept for the next epidemic wave. This parameter is used only when an upward update is triggered and there were very few triggering events during the training data period. Therefore it was estimated directly from data by calculating the correlations between changes over two years and (positive) changes over one year. That is, yearly total numbers of cases were calculated for each country and denoted by $\bar{Y}_i(k) = \sum_{t=52(k-1)}^{52k} y_i(t)$, where $k = 1, ..., 10$ denotes the year. We then formed pairs $\big(\bar{Y}_i(k) - \bar{Y}_i(k-1), \bar{Y}_i(k+1) - \bar{Y}_i(k-1)\big)$ for each country for $k = 2, ..., 9$, and then calculated the regression line for these points including only those for which $\bar{Y}_i(k) - \bar{Y}_i(k-1) > 0$. The regression coefficient 0.756 (raised to power $\alpha_2$) is used as the value for the parameter $\alpha_3$.

Once the parameter has been updated, the last twelve weeks are re-run with the new parameter value. Note that the 4-week ahead projections $\hat{y}(t+\tau|t)$ are only generated when week $t$ is passed for the first time in the algorithm to ensure causality of the projections.

**Downward update.** Unlike for too small $c_i(t)$, there are no clear signs for too high value, except for overshooting projections, in particular, during times of high incidence. Therefore, downward updates are only done on pre-determined times between epidemic waves (that is, in summer). At such time $T$, the performance for the previous year is analysed for all countries. To check for overshooting projections, for each country, we find the weeks from the previous year $T-51, ..., T$ during which the incidence has been on the highest 30th percentile during that year (denote this set of weeks by $\mathbf{T}_i(T)$). The $c_i(t)$ is then reduced by multiplication by a factor

$$\min\left\{1, \left(\frac{\sum_{t\in\mathbf{T}_i(T)} \sum_{\tau=1}^{4} y_i(t+\tau)}{\sum_{t\in\mathbf{T}_i(T)} \sum_{\tau=1}^{4} \hat{y}_i(t+\tau|t)}\right)^{\alpha_2}\right\}. \tag{9}$$

The reduction is done in a piecewise linear way, such that the linear descent takes place on weeks $T+1$ to $T+6$.

##### S4.2 Fitted parameter values

We can run the parameter fitting scheme using the EKF for the different model versions (full network, mean field network, isolated models) and for different tests for interpretability (scaling by population, silencing of certain tuning parameters) and for performance by increasing the data samples using for training; see Main Text for all detailed descriptions. Supplementary Table S1 lists all parameters and their associated equations, while Supplementary Table S2 summarizes all parameter values.

We notice that $\kappa_1$ is quite large. The Kalman updates are therefore mainly focused on $S$ and $I$, while the updates of $\beta$ are fairly small. In addition, $k_1$ is relatively big, strongly driving $\beta$ towards the reference value, in particular with the network models, where the reference value is heavily influenced by the network ($k_2 \approx 0.8$ in all network cases). Another noteworthy issue concerning the parameters is that some parameters are not very well identifiable, which results in relatively large differences for the parameter values when the training is done using the first four years of data or all ten years of data, while the performance did not change considerably. In particular, $\gamma_2$ changes considerably in the network model, indicating that the inclusion of the neighbour network is not very important for forecasting performance.

In this work, we did not intervene on these factors so as to provide as generic a methodology as possible; however, further fine-tuning is possible, *e.g.*, when focusing on particular countries with known peculiarities.

Supplementary Table S1: Summary of the 11 tuning parameters fitted by simulated annealing. The equations marked with MT are in Main Text.

| Group | Symbol | Explanation | Ref. Eq. |
|---|---|---|---|
| Network | $\gamma_1$ | Coefficient for the travel and cross-border work networks | (1) (MT) |
| | $\gamma_2$ | Coefficient for the neighbour network | (1) (MT) |
| State noise | $\kappa_1$ | Coefficient for the Langevin covariance | (4) |
| | $\kappa_2$ | Coefficient for the state-independent noise covariance | (4) |
| Measurement noise | $\rho_1$ | Coefficient for the state-dependent measurement noise | (6) |
| | $\rho_2$ | Coefficient for the state-independent measurement noise | (6) |
| $\beta$-dynamics | $k_1$ | Coefficient for $\beta$-dynamics | (5) |
| | $k_2$ | Coefficient for neighbour average vs. grounding value in $\beta$-dynamics | (5) |
| | $k_3$ | Coefficient for grounding value of $\beta$ | (5) |
| Adaptive $c_i(t)$ estimation | $\alpha_1$ | Coefficient for initial value for $c_i(t)$ | (7) |
| | $\alpha_2$ | Transformation exponent between case numbers and $c_i(t)$ | (8),(9) |

Supplementary Table S2: Fitted parameter values for different models. Note that with the two mean-field models, the first parameter is $\gamma$, and it is not comparable with values of $\gamma_1$. Also the $\gamma_j$ values for the scaled network are not comparable with other network models. When the network is only used in $\beta$-dynamics, $\gamma$ is set to 1, since its precise value would be lost in the normalisation in (5).

| Model | $\gamma$ / $\gamma_1$ | $\gamma_2$ | $\kappa_1$ | $\kappa_2$ | $\rho_1$ | $\rho_2$ | $k_1$ | $k_2$ | $k_3$ | $\alpha_1$ | $\alpha_2$ |
|---|---|---|---|---|---|---|---|---|---|---|---|
| Factor | 1 | 1 | 1 | $10^{-8}$ | $10^{-2}$ | $10^{-6}$ | $10^{-1}$ | 1 | 1 | 1 | 1 |
| Network | 1.787 | 0.666 | 2257 | 1.032 | 1.19 | 4.074 | 1.71 | 0.832 | 1.325 | 0.812 | 1.442 |
| Isolated | 0 | 0 | 5506 | 79.63 | 6.65 | 28.44 | 0.33 | 0 | 1.393 | 0.670 | 0.794 |
| Mean-field | 6.246 | 0 | 4597 | 20.37 | 1.28 | 10.35 | 0.90 | 0.766 | 1.194 | 0.809 | 1.568 |
| Scaled network | 2.943 | 0.570 | 3496 | 2.008 | 3.76 | 8.178 | 1.15 | 0.801 | 1.260 | 0.817 | 1.514 |
| Case $k_2 = 0$ | 13.36 | 3.605 | 8233 | 8.894 | 10.5 | 35.99 | 0.198 | 0 | 1.086 | 0.641 | 0.975 |
| Mean-field in $\beta$ | 1 | 0 | 2186 | 49.84 | 0.75 | 6.365 | 1.10 | 0.858 | 1.433 | 0.642 | 1.038 |
| Network, full data | 1.754 | 0.064 | 2807 | 0.4249 | 0.95 | 7.506 | 2.17 | 0.846 | 1.312 | 0.884 | 0.944 |
| Isolated, full data | 0 | 0 | 3922 | 62.12 | 4.95 | 26.26 | 0.443 | 0 | 1.398 | 0.807 | 0.913 |

### S5 Performance evaluation

For performance evaluation, we define the $\tau$-week ahead forecasts $\hat{y}_i(t+\tau|t) = c_i(t+\tau)\hat{I}_i(t+\tau|t)$ where $\hat{I}_i(t+\tau|t)$ is obtained by simulating the noise-free model forward in time for $\tau$ weeks, starting from the Kalman filter state estimate after the update on week $t$.

The total forecasting error is defined for the $i$-th country by

$$E_i(\mathbf{T}) = \sum_{\tau=1}^{4} \sum_{t+\tau\in\mathbf{T}} \left|\sqrt{y_i(t+\tau)} - \sqrt{\hat{y}_i(t+\tau|t)}\right| .$$

Here, $\mathbf{T}$ denotes the time interval or a collection of time intervals used for evaluation. The square root is used as transformation to stabilize variance, so as to balance the performance metric between times of high and low incidence, and between large and small countries. Note that weeks $t$ where data for country $i$ are missing for weeks $t-3, ..., t$, are excluded from $E_i$.

The sum $\sum_{i=1}^{28} E_i$ is also used as the cost function for parameter fitting, using the first four years $\mathbf{T} = \{1, ..., 202\}$ as the training data (until week 32 in 2018, see Figure 2). The remaining data are used as test data $\mathbf{T} = \{203, ..., 499\}$.

As a reference to benchmark our predictions, we simply use the latest data point as a forecast for the next four weeks:

$$E_i^{(\mathrm{ref})}(\mathbf{T}) = \sum_{\tau=1}^{4} \sum_{t+\tau\in\mathbf{T}} \left|\sqrt{y_i(t+\tau)} - \sqrt{\bar{y}_i(t)}\right|$$

where $\bar{y}_i(t) = y_i(t)$ if these data exist, or equals the value of the previous existing data point, if $y_i(t)$ does not exist. We measure how much all different models, used for evaluation, improve the overall predictions w.r.t. the reference forecast, that is,

$$\text{Performance gain} = 1 - \frac{\sum_{i=1}^{28} E_i(\mathbf{T})}{\sum_{i=1}^{28} E_i^{(\mathrm{ref})}(\mathbf{T})}\,, \tag{10}$$

calculated separately for the training data period and the test data period. In addition, to evaluate performance during the wave onset and peak periods, the errors $E_j$ are collected over time intervals covering the onsets of all epidemic waves (except for 2020–2022, which were heavily affected by the COVID-19 pandemic). Here, we no longer considered the split between training and testing data. These time intervals were determined by identifying weeks $W$ when the European-wide incidence crossed 120, and taking the interval from $W-8$ to $W+1$. Additionally, we collect all time intervals around the peak incidence, identified by weeks $\hat{W}$ with the highest incidence during each epidemic season, and take the interval $\hat{W}-5$ to $\hat{W}+4$ (see Figure 2).

#### S6 Calibration for probabilistic forecasts

As mentioned above, if the covariance matrices $Q$ and $U$ and the initial uncertainty covariance $P_0$ are multiplied by a constant, it has no effect on the Kalman filter state estimate, and hence the cost function used for fitting the model parameters is oblivious to such scaling. However, covariance scaling has an effect on the uncertainty of the state estimation, which is important for probabilistic forecasts. Therefore, an additional calibration step is required after fitting other parameters in order to match the forecasting uncertainties to actual errors. The best way to produce probabilistic forecasts is to simulate trajectories from the full stochastic model and gather statistics from these simulations. For these stochastic simulations, we introduce a coefficient $K$ to control the uncertainty in the stochastic projections. To generate probabilistic forecasts, we simulate 1000 trajectories forward in time. These trajectories are initialised from a random initial state drawn from $\mathcal{N}\big(\hat{x}(t), K^2 P_t\big)$. Trajectories are then simulated forward in time using the stochastic model (4)–(5), where all noise terms $w_j$ for $j = 1, ..., 5$ are scaled with $K$. Both state-dependent and state-independent measurement noise corresponding to (6), again multiplied by $K$, is then added to the projected case numbers.

To find a good value for $K$, we generated 4-week ahead probabilistic forecasts on 58 different weeks covering the last two epidemic seasons (every week during high incidence and every second week during low incidence). For the probabilistic forecasts, we generated several different percentiles for the projections, and then calculated the share of cases where the real incidence (for forecast weeks 1–4) was above each percentile. This was repeated with several different values of $K$ and the results are shown in Supplementary figure S2.

Due to nonlinear dynamics, the median of the 1000 trajectories does not coincide with the noise-free projection ($K = 0$) that was used for tuning the method. To account for potential bias, we also tried a scheme where all the simulated percentiles were adjusted by adding the difference of the noise-free projection and the median. From Supplementary figure S2, it can be observed that this adjustment indeed helps with mitigating bias in the forecasts. Therefore, we will choose the adjusted forecasts with $K = 2.25$ to be used for probabilistic forecasts.

#### S7 Interpretability of the model

Here, we report the detailed results of the interpretability tests described in the Main Text. In particular, Figure S3 shows the performance gain for all tests conducted, upon training and test data, as well as sensitive times such as wave onset and incidence peaks (compare with Figure 4 in the Main Text).

The scaled network is obtained by scaling the rows of $G^{(1)} + G^{(2)}$ and $G^{(3)}$ to a fixed percentage of the populations of the respective countries, in order to see the effect of population sizes in prediction performance: are predictions on more (or less) populated countries more precise? The change is not significant, meaning that networks built directly with population data already perform well, without resorting to scaling.

Setting $k_2 = 0$ means excluding network effects from the dynamics of the transmission parameter $\beta$. We observe a decrease in performance, which highlights that transmission is significantly influenced by neighbouring countries, and its dynamics should account for these effects to improve predictions.

The "mean-field $\beta$-dynamics" refers to when network effects only affect the $\beta$-parameter dynamics, while the S-I dynamics is ruled by isolated models. This leads to an almost equivalent performance to the full mean-field model.

Finally, we use all data as training data, to see whether augmenting significantly the training period improves the model performance. We do not observe significant improvements given by more data being processed for training, highlighting the parsimony and effectiveness of our models.

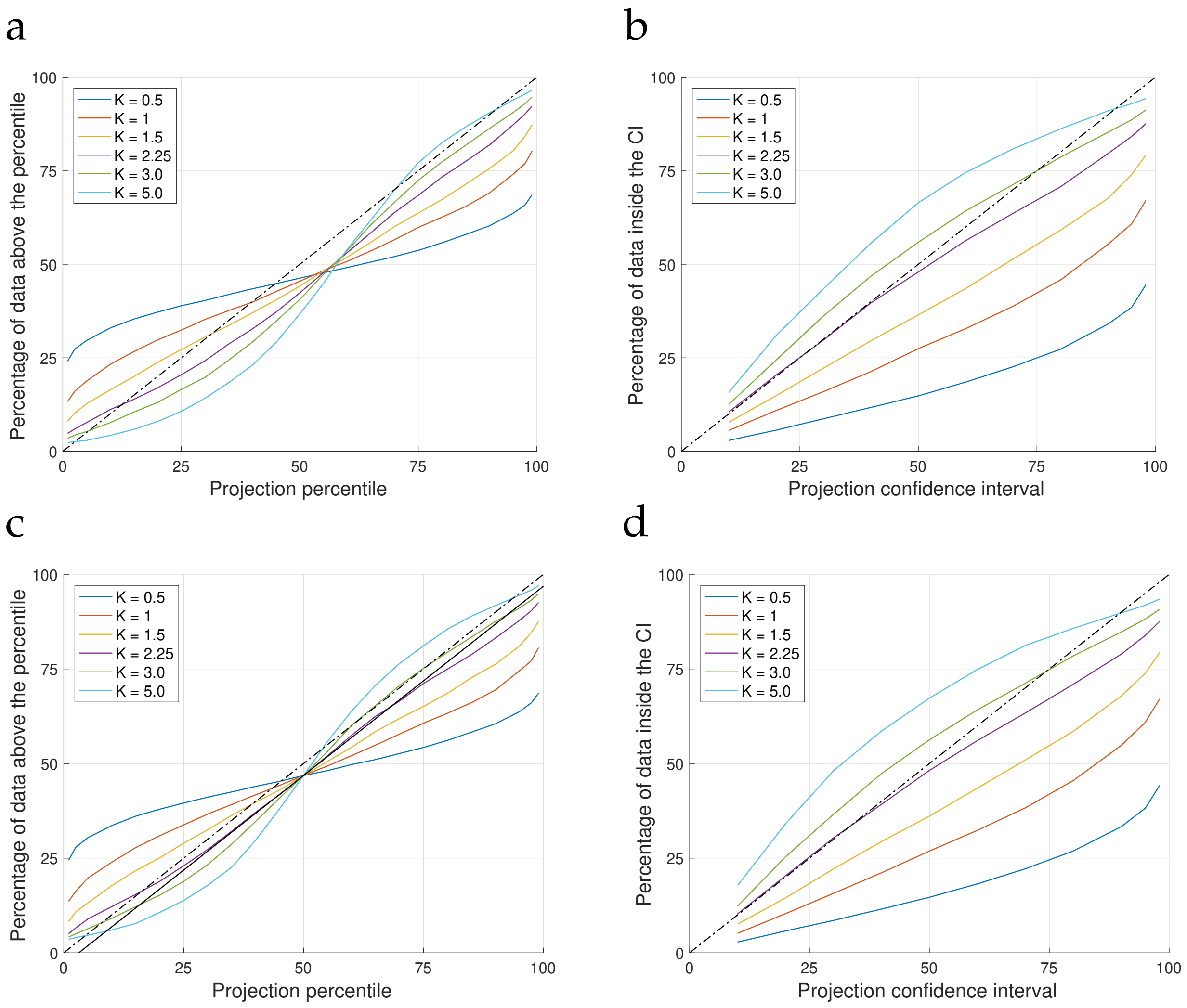


Supplementary Figure S2: Calibration results. In all plots, the black dash-dotted line indicates a perfectly calibrated (and unbiased in panels a and c) forecast. **a:** Share of cases when the true incidence is above different forecast percentiles for the non-adjusted forecasts. With different tested values of $K$, the true incidence is above the forecast median (Projection percentile 50) in 36.7–46.3% of forecasts, indicating some bias (underestimation) in the forecasts. **b:** Share of cases when the true incidence is inside different confidence intervals for the non-adjusted forecasts. **c:** Share of cases when the true incidence is above different forecast percentiles for the adjusted forecasts. Due to the adjustment, all curves coincide at the median, and 46.8% of true incidence is above the forecast median, which is a clear improvement on the non-adjusted forecasts, in particular for appropriate values of $K$. The black solid line is parallel to the line corresponding to perfect calibration, but it is offset to pass through the forecast median point. The case with $K = 2.25$ matches this line at best (also $K = 2$ and $K = 2.5$ were tested but are not shown). **d:** Share of cases when the true incidence is inside different confidence intervals for the adjusted forecasts.

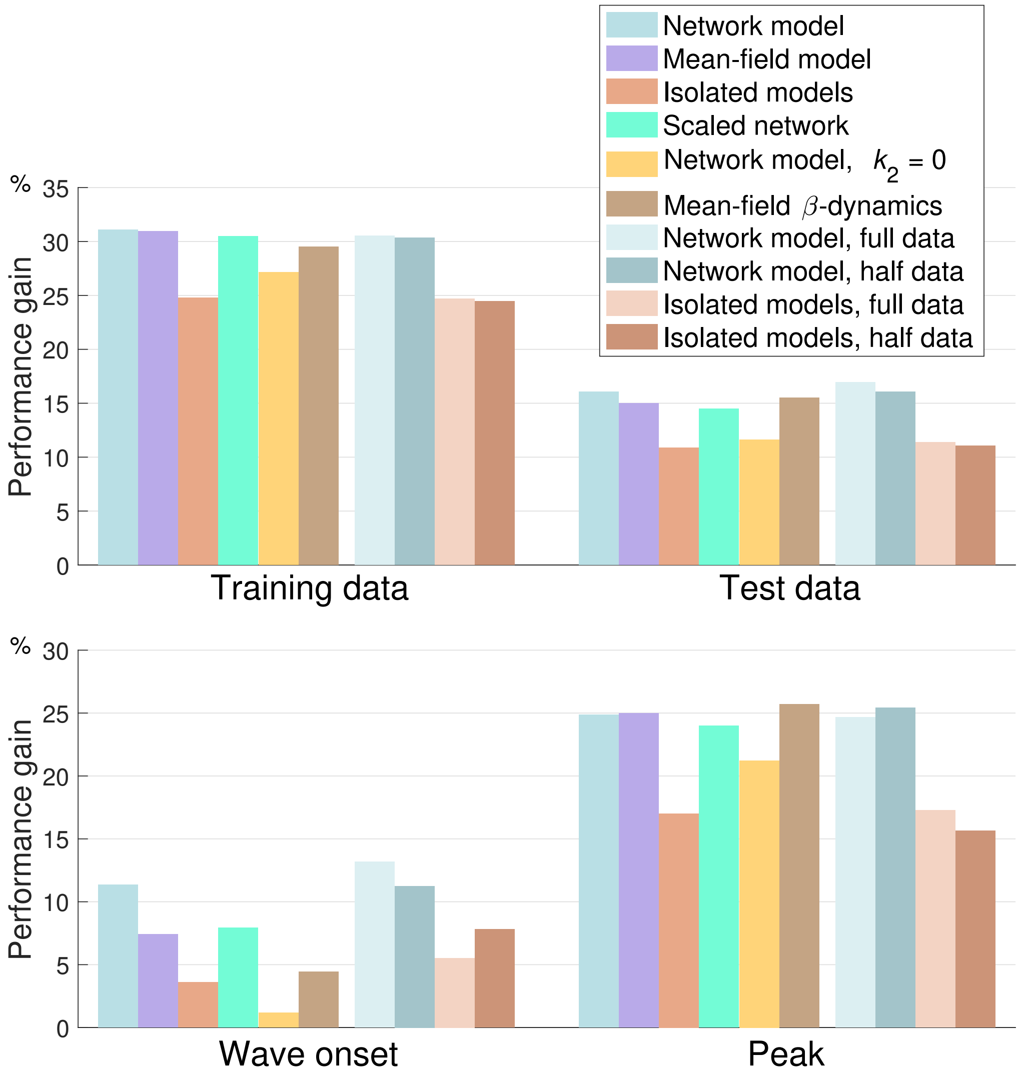


Supplementary Figure S3: Performance for the supplementary experiments for interpretability. The first three bars correspond to the results shown in Figure 4. The scaled network is obtained by scaling the rows of $G^{(1)} + G^{(2)}$ and $G^{(3)}$ to a fixed percentage of the populations of the respective countries. Setting $k_2 = 0$ removes the effect of the network in the $\beta$-parameter dynamics. The "Mean-field $\beta$-dynamics" refers to a model where the mean-field network is used, but only in $\beta$-parameter dynamics, while the calculation of $F_{S_i \to I_i}$ is done as with isolated models. The last four bars are obtained by using either all ten years of data for parameter tuning, or only the first two years. The evaluations shown here are still done using the original split between training and testing sets.
